# Usability and performance of the MicroGEM Sal6830™, an RT-PCR saliva-based point-of-care platform to detect SARS-CoV-2 in primary healthcare settings with non-laboratory trained operators

**DOI:** 10.1101/2023.01.01.22283267

**Authors:** Vanessa Mills, Teresa Owens Tyson, Rachel Helton, Paula Hill-Collins, Sarah Hubbard, Anchi Scott, Jeff A. Hickey, Brian E. Root, Rory O’Brien, Jillian Conte, Jo-Ann L. Stanton, Jeff D. Chapman

## Abstract

The beginning of the COVID-19 pandemic demonstrated how few point-of-care diagnostic tools were available that could be safely and easily operated by healthcare workers with no laboratory training. The gold-standard test, and initially the only test, used RT-PCR with nasal pharyngeal swabs (NPS). Two issues quickly emerged: 1) RT-PCR required central laboratory processing leading to significant time delays and 2) NPS collection causes discomfort, is inappropriate for ongoing repeat sampling of individuals (e.g., frontline healthcare workers) and poses difficulty when obtaining samples from some sections of the population (e.g. some elderly and young children). The Sal6830™ platform is a fully self-contained, RT-PCR point-of-care device for detecting SARS-CoV-2 from saliva that takes less than thirty minutes to complete. In this study we tested the usability of the Sal6830™ platform by healthcare workers unfamiliar with the instrument at two community clinics: Care 4 U Community Health Center (Miami, Florida, USA) and St. Mary’s Health Wagon (Wise, Virginia, USA). Staff participated in three tests: 1) determining SARS-CoV-2 status from blinded positive and negative saliva samples, 2) a clinical study comparing SARS-CoV-2 detection with a comparator point-of-care technology from the same patient and 3) completing a survey designed to measure comfort and confidence using the Sal6830™ point-of-care device having received no training. Participants overwhelming found the Sal6830™ platform easy and intuitive to use, successfully called SARS-CoV-2 status of contrived, blinded samples and measured a 93.3% overall percent agreement when comparing patient samples across two point-of-care technologies.

## Introduction

COVID-19 spread rapidly from first detection of SARS-CoV-2 in late 2019 to the declaration of a pandemic by March 2020. The gold standard to diagnose and track the virus is reverse transcriptase polymerase chain reaction (RT-PCR), a highly accurate test but, at the time, anchored to central laboratory facilities in the hands of highly trained specialists [1]. Turn-around times on test results could be as long as five days leading to patient loss-to-follow-up, increased opportunity for viral spread, and the unnecessary quarantine of uninfected people. In addition, the available tests required nasopharyngeal swab (NPS) samples that increase medical professionals’ exposure to potentially contagious patients, caused patient discomfort, and contributed to inadequate sampling leading to inconsistent results [2–4]. For certain patient groups, such as pediatric patients and patients with mental disabilities, collection of NPS samples can be difficult or unattainable [3].

One solution that offered to overcome some of these challenges was to test patients at the point-of-care (POC) in real time in order to reduce wait time for results. However, early in the pandemic, technologies for RT-PCR that could be operated by non-laboratory trained healthcare workers at the POC were limited. There were some RT-PCR POC platforms available early in the pandemic, such as the Xpert Xpress SARS-CoV-2 test (Cepheid) and the cobas® SARS-CoV-2 & Influenza A/B Nucleic Acid Test for use on the cobas® Liat System (Roche Molecular Systems, Inc.) [5]. These technologies utilized eluate from a healthcare provider-collected or instructed NPS placed in a variety of viral transport media. In these systems a subsample of the eluate is transferred to an assay-specific detection platform where the sample is processed and undergoes RT-PCR; this additional sample handling introduces the opportunity for cross-contamination between different samples or patient ID mix up. Also, the operator may be exposed to a potentially hazardous infectious material during sample transfer. The off-platform sample handling can also present difficulty for non-laboratory trained individuals to operate safely with confidence.

It was determined early in the pandemic that SARS-CoV-2 can be detected in a variety of biological sample types [6]. Saliva can be collected via swabbing or from stimulated or unstimulated manipulation of the salivary gland and provides non-invasive, lower cost biospecimen collection [7]. SARS-CoV-2 can be detected reliably from saliva [7]. Connor *et al*. found in their comparative study that substituting a saliva sample for NPS in a laboratory based SARS-CoV-2 RT-PCR assay without adjusting for sample-type, was acceptable for diagnostic testing, particularly when required for repeat-testing or working with challenging patients [3]. Tarantini *et al*. also noted that in addition to patient discomfort when NPS are collected, incorrect application of the swab can mean the nasopharynx target site is not reached [8] with a significant negative impact upon test reproducibility, reliability, and standardisation. The skill of the sample collector does not similarly impact self-collection of a saliva sample.

Even after multiple waves of SARS-CoV-2 outbreaks around the world and many more POC *in vitro* diagnostic devices issued with emergency use authorization by the U.S. Food and Drug Administration, a call for a saliva-based, RT-PCR, POC device has not received a good response [9]. The MicroGEM Sal6830™ SARS-CoV-2 Saliva Test is the first such test with emergency use authorization to respond to this call and in doing so to overcome many of the challenges described here. The Sal6830™ uses saliva that the patient self-deposits directly into the all-in-one processing cartridge which they cap/seal themselves, protecting the operator from handling a potential biohazard and, as the test is run immediately, significantly reducing the possibility of patient sample mix up. Figure 1 illustrates the processing of COVID-19 saliva samples using the Sal6830™ system. The Sal6830™ cartridge chemistry has been optimised for saliva and a bead-based viral capture system traps and concentrates intact virions rather than viral debris or un-encapsulated viral RNA, potentially differentiating between infectious and non-infectious patients.

**Fig 1:**
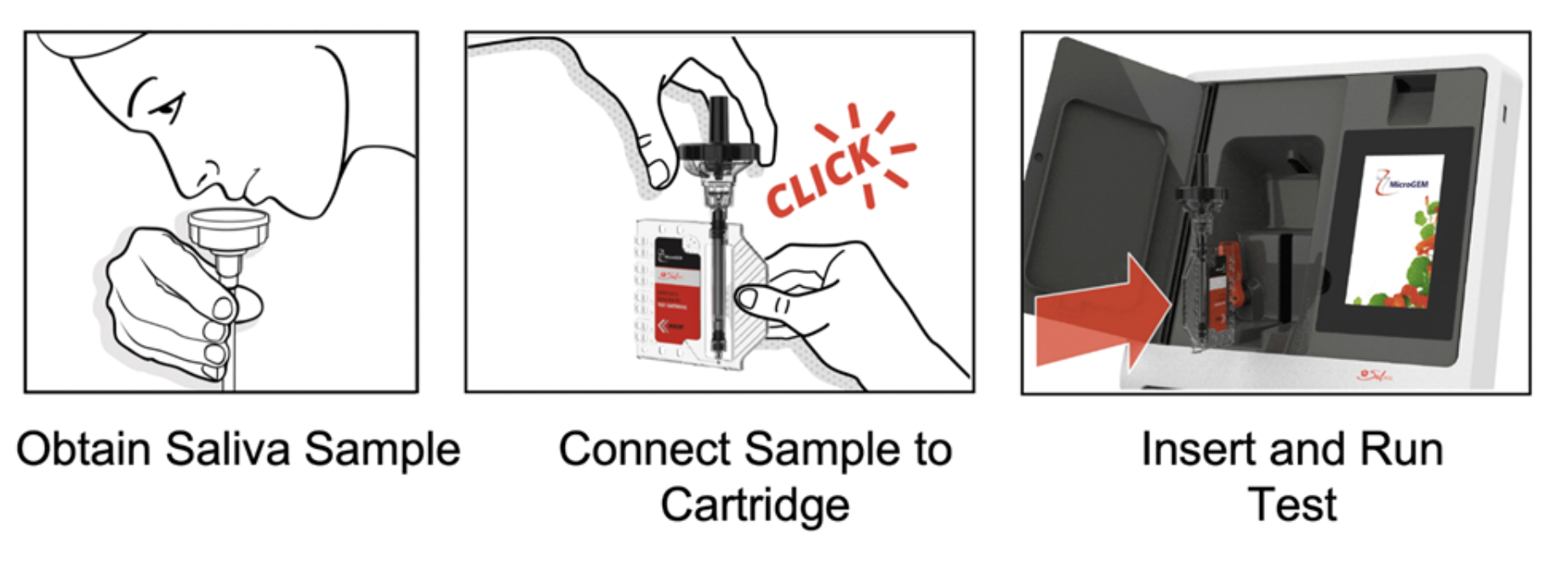
Sample collection and test run using Sal6830™. The patient deposits approximately 1 ml of saliva directly into the Sal6830™ collection cup and twist-seals the cap in place to permanently contain the sample within the extraction tube assembly. The act of twisting the cap releases viral capture reagent into the sample which is then gently swirled to mix before being placed into the test cartridge and slotted into the Sal6830™ instrument. The run is initiated via a touch screen and a result (Detected, Not Detected or Invalid) is displayed on the screen after approximately thirty minutes. The spent cartridge is removed from the device and deposited directly into biohazard waste.

The ability to rapidly triage patients as they present to the clinic (i.e. the POC) enables healthcare workers to immediately provide proper care and to direct resources appropriately. Prompt triage means that treatments can be administered earlier, infected and non-infected patients can be rapidly identified, and isolated as needed, and valuable personal protective equipment can be prioritised for use with those patients known to have COVID-19 infection. However, a POC approach can only be successfully implemented to maintain patient flow if healthcare workers are confident to use it and can demonstrate that the results they generate from the platform are accurate. In this study, we investigated whether healthcare workers without previous laboratory training were able to use the Sal6830™ system with confidence and accuracy to detect patients with COVID-19 at the POC.

## Materials and Methods

### Health clinics and ethical approval

Clinical performance and usability studies were performed at two centres: Care 4 U Community Health Center (Miami, Florida, USA) and St Mary’s Health Wagon (Wise, Virginia, USA). Ethical approval for these studies was granted under clinic Institutional Review Board (IRB) Study Numbers 1313475 and 1315538, respectively. The IRB Tracking Number for this study was 2021278. The locations and highest academic qualification of all participating healthcare workers are provided in Table 1. Saliva used for blind samples was collected under WCG IRB study number 1294772.

**Table 1.** Education and title of non-laboratory trained healthcare workers.

| OPERATOR | TITLE | DEGREES/ASSOCIATIONS | LOCATION |
| --- | --- | --- | --- |
| A | Medical Assistant | High School Diploma | St Mary's Health Wagon |
| B | Nurse | Licensed Practical Nurse | St Mary's Health Wagon |
| 1 | Medical Assistant | High School Diploma | Care 4 U |
| 2 | Medical Assistant | High School Diploma | Care 4 U |
| 3 | Nurse | MSc. Nurse Practitioner | Care 4 U |
| 4 | Medical Assistant | High School Diploma | Care 4 U |
| 5 | Family Nurse Practitioner | MSc Nursing | Care 4 U |
| 6 | Medical Assistant | BA | Care 4 U |
| 7 | Chief Operating Officer | RN, BSN, MPH | Care 4 U |

### Preparation of blinded samples for clinical tests

Verified negative saliva was collected from donors and pooled. A SARS-CoV-2 gamma-irradiated standard supplied by the US Centre for Disease Control and Prevention (BEI Resources, Manassas, VA, USA, catalogue number NR52287, lot number 70039068) was obtained at a viral stock concentration of 7.65 × 10^8^ genome equivalents (GE)/ml according to the certificate of analysis. The verified negative pooled saliva was stored at -80°C until blinded samples were prepared. Blind positive samples were prepared by diluting the concentrated viral stock to a final concentration of 1.9 × Limit of Detection (LoD) for Sal6830™, spiked into the verified negative pooled saliva. Blind positive samples were prepared as 1.1 mL aliquots. Blind negative samples were prepared as 1.1 mL aliquots of the verified negative pooled saliva. Blinded samples were stored at -80°C until required. Each sample was labeled with a unique alphanumeric code known only to MicroGEM’s clinical trial director (N = 48). The blinded samples were shipped by overnight courier to the participating clinics. Six non-laboratory trained healthcare workers with no previous experience using Sal6830™ performed SARS-CoV-2 detection on the blinded samples according to the Sal6830™ operating instructions. Further details of sample preparation are provided in the Supporting Information.

### Clinical performance evaluation of Sal6830™

The performance of the Sal6830™ SARS-CoV-2 Saliva Test at POC was evaluated in a prospective multisite clinical study. A flow chart of the study design is illustrated in Fig 2. Study participants were consented into the study at single presentation to the clinic. All participants were screened against the inclusion/exclusion criteria, gave their informed consent, and provided a baseline survey of subject demographics and COVID-19 status. The study population included individuals aged 5 years and over. Pregnant women could participate in the study. Individuals were included if their healthcare provider suspected COVID-19 and they were able to provide consent. Exclusion criteria included an inability to tolerate or provide two samples, involvement in planning and/or conducting the study, or previous participation in a Sal6830™ SARS-CoV-2 Saliva Test study. Further details for consenting participants are available as part of the Supporting Information.

**Fig 2.**
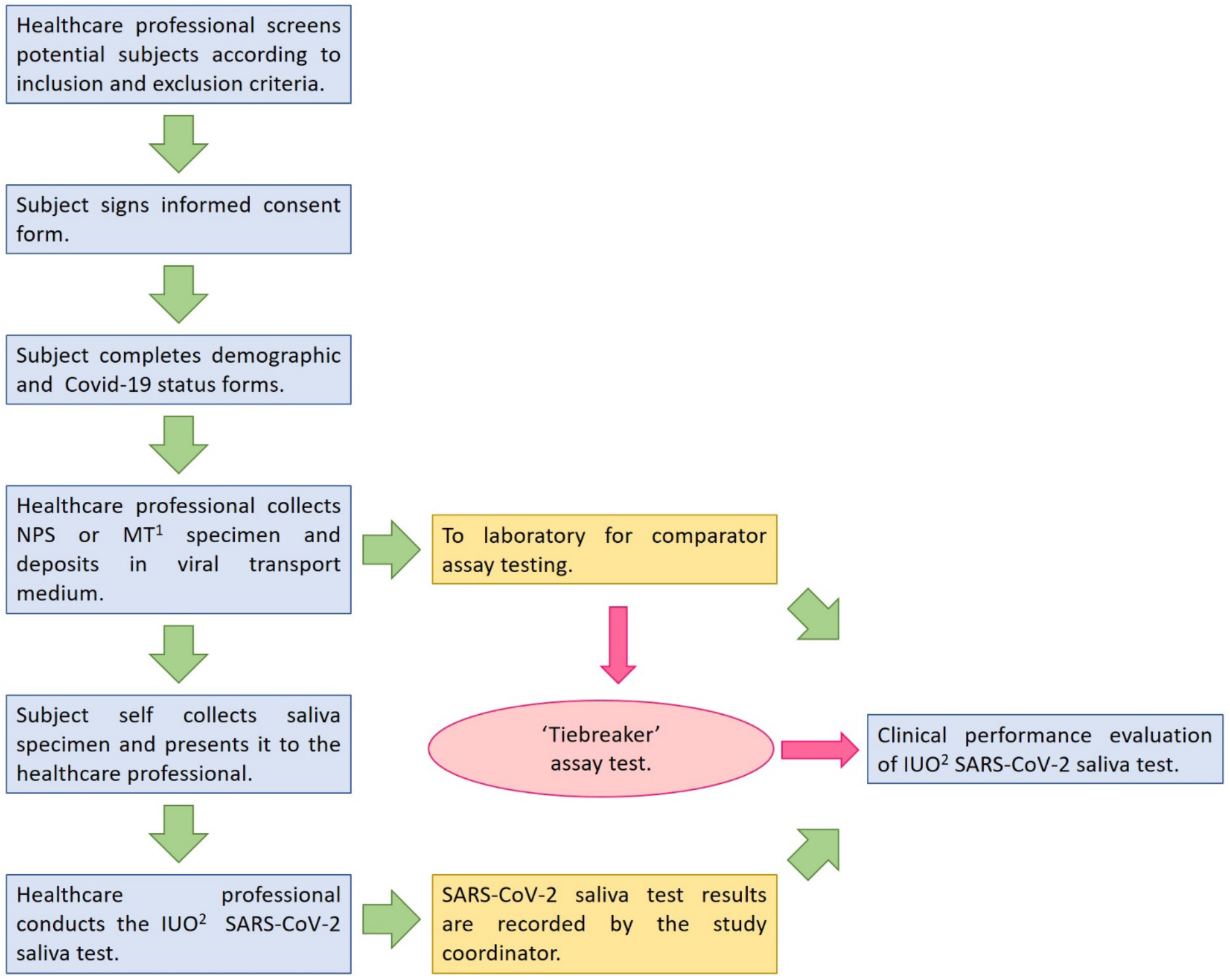
General Study Design for Sal6830™ clinical comparison. A Tiebreaker test was performed by running a second Cepheid Xpert Xpress CoV-2/Flu/RSV assay if results were discordant. ^1^ MT, Mid-Turbinate; ^2^ IUO, Informational Use Only.

Each participant provided two samples. The first sample was a NPS that was shipped to either Obstetrical Associates Laboratory Services, Fall River, Massachusetts, USA or Intermountain Laboratory Services, Murray, Utah, USA for testing on the comparator POC platform. The comparator platform chosen for this study was the Cepheid Xpert Xpress CoV-2/Flu/RSV operated in accordance with the manufacturer’s instructions. The second sample was a patient collected saliva sample for processing on the Sal6830™ as per the instructions for use (IFU). Patients self-deposited a saliva sample directly into the Sal6830™ extraction tube assembly eliminating any additional sample handling by the operator. The same six non-laboratory trained operators that performed the blind sample evaluation performed all prospective clinical tests undertaken as part of this evaluation.

### Sal6830™ usability questionnaire for point-of-care sites

The confidence and comfort of non-laboratory staff using the Sal6830™ was assessed through a usability questionnaire (Likert Scale). The questionnaire was completed by the six non-laboratory healthcare workers who took part in the other experiments and an additional three non-laboratory staff who also ran the Sal6830™ platform at Care 4 U Community Health Center on patients not recruited into the formal study. A copy of the detailed data are provided in the Supporting Information.

## Results and Discussion

Results obtained from blinded samples processed on the Sal6830™ platform operated by non-laboratory trained healthcare workers are detailed in Table 2. All results of the negative saliva samples and positive samples were concordant with the expected results. One positive sample returned an invalid result, which is not a discordant result. Per the IFU in a real world setting, this patient would have been requested to resubmit a sample for testing rather than being cleared of COVID-19 infection. These results suggest that the Sal6830™ can be run safely and correctly by healthcare workers without training in laboratory practice and that the operation of the Sal6830™ platform is straightforward and intuitive.

**Table 2.** Operator performance of Sal6830™ with blinded samples at 1.9 × LoD at two clinical sites.

| SITE & OPERATOR | BLINDED PANEL TESTING RESULTS |  |  |  |  |  |
| --- | --- | --- | --- | --- | --- | --- |
|  | Positive Samples |  |  | Negative samples |  |  |
|  | Number Tested | Pos. | Neg. | Number Tested | Pos. | Neg. |
| CARE 4 U TOTALS | 12 | 12 | 0 | 12 | 0 | 12 |
| 1 | 3 | 3 | 0 | 3 | 0 | 3 |
| 2 | 3 | 3 | 0 | 3 | 0 | 3 |
| 3 | 3 | 3 | 0 | 3 | 0 | 3 |
| 4 | 3 | 3 | 0 | 3 | 0 | 3 |
| ST MARY'S<br>HEALTH WAGON<br>TOTALS | 12 | 11 | 0 | 12 | 0 | 12 |
| A | 6 | 5 | 0 | 6 | 0 | 6 |
| **B** | 6 | 6 | 0 | 6 | 0 | 6 |
| STUDY TOTAL | 24 | 23 | 0 | 24 | 0 | 24 |

Sal6830™ clinical performance in the hands of non-laboratory trained healthcare workers was tested in a comparator, prospective trial. The same six healthcare workers who had processed the blinded samples carried out this work. Patients suspected of having COVID-19 were recruited on presentation at the clinic. They provided two samples, one tested using the Sal6830™ and the second using a different, RT-PCR based POC platform (Cepheid Xpert Xpress CoV-2/Flu/RSV test). Critical differences between the two tests are the sample type (Sal6830™ uses saliva whereas the Cepheid Xpert Xpress CoV-2/Flu/RSV requires a NPS sample) and the degree of sample handing by the operator when running the test. Patients collected their own saliva sample directly into the Sal6830™ extraction tube assembly (‘test cartridge’) while the NPS required further steps to transfer swab eluate in viral transport medium to the Cepheid detection cartridge. Table 3 details the clinical performance data for both tests. The positive percent agreement (PPA), negative percent agreement (NPA), and overall percent agreement for the clinical study were 87.2% (Confidence Interval (CI) = 74.8% - 94.0%), 97.2% (CI = 90.4% - 99.2%) and 93.3% (CI = 88.6% - 98.1%), respectively. These high levels of agreement between results obtained by non-laboratory trained healthcare workers and those from central laboratory facilities confirm that Sal6830™ performed according to expectations in a POC setting when operated by untrained, lay operators.

**Table 3.** Clinical performance of Sal6830™ SARS-CoV-2 saliva test.

| Combined Sites* |  | Cepheid Xpert Xpress CoV-2/Flu/RSV |  |  |
| --- | --- | --- | --- | --- |
|  |  | Positive | Negative | Total |
| Sal6830™ | Positive | 41 | 2 | 43 |
|  | Negative | 6 | 70 | 76 |
|  | Total | 47 | 72 | 119 |

\*PPA = TP/(TP+FP); NPA = TN/(TF+FP); TP = True Positive; TN = True Negative; FP = False Positive; FN = False Negative.

The predictive agreement between the Cepheid and Sal6830™ POC platforms were similar but not identical. We attribute this to underlying differences in how the two platforms capture SARS-CoV-2 viral material. By processing whole NPS eluate the Cepheid detects both viral particles and associated but non-infectious viral debris [10]. The Sal6830™ viral capture chemistry only detects intact viral particles that are more likely to be infectious. Detection differences between the two sample types were recently reviewed in McPhillip and MacSharry (2022) [11] with saliva from the same patient shown to be positive when NPS is negative, and vice versa.

A summary of healthcare workers responses to the usability questionnaire is provided in Table 4. All healthcare workers participating in this trial found the Sal6830™ easy to use and they did not require additional technical support. The system and test functions were well integrated, with test results easy to interpret. Overall, it was found that healthcare professionals would learn to use the Sal6830™ quickly and in the clinics participating in this trial, all users were confident they could conduct the test. What is more, they planned to use Sal6830™ frequently in POC settings. This confirms healthcare workers were comfortable and confident using Sal6830™ to process patient samples in a POC setting without technical backup.

**Table 4.** Average response to System Usability Questionnaire for Sal6830™. Values were recorded as 1=strongly disagree. 5=strongly agree (N = 9).

| Question | Average response ± SD* |
| --- | --- |
| The MicroGEM Sal6830™ SARS-CoV-2 Saliva test works well in Point-of-Care settings. | 4.7 ± 0.47 |
| I found the MicroGEM Sal6830™ SARS-CoV-2 Saliva test was unnecessarily complex. | 1.2 ± 0.42 |
| I found the MicroGEM Sal6830™ SARS-CoV-2 Saliva test was easy to use. | 4.6 ± 0.68 |
| I will need the support of a technical person to be able to use the MicroGEM Sal6830™ SARS-CoV-2 Saliva test. | 1.1 ± 0.31 |
| I found the various functions in the MicroGEM Sal6830™ SARS-CoV-2 Saliva test well integrated. | 4.7 ± 0.47 |
| I would imagine that most healthcare professionals would learn to use the MicroGEM Sal6830™ SARS-CoV-2 Saliva test quickly. | 4.7 ± 0.47 |
| I found the MicroGEM Sal6830™ SARS-CoV-2 Saliva test difficult to use. | 1.1 ± 0.31 |
| I felt confident using the MicroGEM Sal6830™ SARS-CoV-2 Saliva test. | 4.9 ± 0.31 |
| I needed to learn a lot of things before I could get going with the MicroGEM Sal6830™ SARS-CoV-2 Saliva test. | 1.8 ± 1.31 |
| I found the MicroGEM Sal6830™ SARS-CoV-2 Saliva test results easy to interpret. | 5 ± 0 |
| Would you recommend this device to another (non-laboratory) healthcare professional. | 8 participants answered 'Definitely yes'. 1 said 'Probably yes' |

\*Standard deviation shows population variance.

## Conclusion

In this report we demonstrate that the Sal6830™ SARS-CoV-2 saliva test can be used accurately and easily by non-laboratory trained healthcare workers requiring no technical support or user training. Study participants felt confident to use the Sal6830™ with patient samples and were able to correctly deliver test results. The Sal6830™ platform returned results equivalent to the Cepheid GeneXpert (93.3% overall percent agreement) but was easier to use with no requirement for additional sample handling or for a nurse-collected NPS. Rather, the Sal6830™ used a simple saliva sample that was easily and painlessly self-supplied by the patient. Sal6830™ uses the gold-standard RT-PCR viral detection technology but is fully compatible with POC settings with non-laboratory trained healthcare workers able to operate the system with confidence.

## Data Availability

All data produced in the present study are available upon reasonable request to the authors

## Acknowledgements

We thank the healthcare workers and participants who took part in this study for their generosity in providing their samples and feedback.

