## Supporting Information for "Usability and performance of the MicroGEM Sal6830™, an RT-PCR saliva-based point-of-care platform to detect SARS-CoV-2 in primary healthcare settings with non-laboratory trained operators"

V Mills, et al (2023).

### Supporting information

#### 1) **LoD for Sal6830™ and details from MicroGEM protocol MSOP067**

The analytical sensitivity (limit of detection or LoD) of the Sal6830™ SARS-CoV-2 Saliva Test was determined by testing serial half-log dilutions of pooled negative saliva specimens spiked with gamma-irradiated SARS-CoV-2 virus (USA-WA1/2020; BEI Resources catalog number NR-52287, Lot number 70039068).

Briefly, the starting titer of the virus stock was $7.65 \times 10^8$ genome equivalents (GE)/mL as obtained from the certificate of analysis from BEI resources and was serially diluted to 765,000 GE/mL. From this dilution a further serial dilution was performed to 498,000 GE/mL. Serial half-log dilutions were then performed down to 50 GE/ml. Dilutions were made from pooled human saliva negative for SARS-CoV-2. Five replicates of each dilution were tested using one lot (1V100375) of the Sal6830™ SARS-CoV-2 Saliva test (**Table S1**). The LoD was estimated to be 6,400 GE/ml using probit analysis and verified by testing an additional 20 replicates with the same lot (**Table S2**). To account for system variability, the LoD study was performed across 16 different Sal6830™ systems and 6 different operators.

**Table S1.** Percent positivity of Sal6830™ SARS-CoV-2 Saliva Test at different viral concentrations in clinical matrix.

| SARS-Co-V-2 (GE/mL) | System Results | | | Detected Chamber Results | | | | Gene Detection | |
|---|---|---|---|---|---|---|---|---|---|
| | Positive | Negative | % Positive | CH5 | CH6 | CH7 | CH8 | E | N |
| 498,561 | 5 | 0 | 100% | 5/5 | 5/5 | 5/5 | 5/5 | 5/5 | 5/5 |
| 157,773 | 5 | 0 | 100% | 5/5 | 5/5 | 5/5 | 5/5 | 5/5 | 5/5 |
| 49,928 | 5 | 0 | 100% | 5/5 | 5/5 | 5/5 | 5/5 | 5/5 | 5/5 |
| 15,800 | 5 | 0 | 100% | 4/5 | 5/5 | 5/5 | 5/5 | 5/5 | 5/5 |
| 5,000 | 5 | 0 | 100% | 4/5 | 2/5 | 3/5 | 5/5 | 4/5 | 5/5 |
| 1,582 | 4 | 1 | 80% | 1/5 | 0/5 | 2/5 | 3/5 | 1/5 | 4/5 |
| 501 | 0 | 3 | 0% | 0/3 | 0/3 | 0/3 | 0/3 | 0/3 | 0/3 |
| 159 | 1 | 3 | 25% | 0/4 | 0/4 | 1/4 | 0/4 | 0/4 | 1/4 |
| 50 | 0 | 5 | 0% | 0/5 | 0/5 | 0/5 | 0/5 | 0/5 | 0/5 |
| 0 | 0 | 5 | 0% | 0/5 | 0/5 | 0/5 | 0/5 | 0/5 | 0/5 |

**Table S2.** Confirmation of LoD of Sal6830™ SARS-CoV-2 Saliva Test

| SARS-Co-V-2 (GE/mL) | System Results | | |
|---|---|---|---|
| | Positive | Negative | % Positive |
| 6,400 | 20/20 | 0 /20 | 100% |

**2) Inclusion and exclusion criteria for participation in the prospective clinical study.**

Subjects were included or excluded from the study following the criteria below:

<u>Inclusion Criteria</u>

- Individuals suspected of SARS-CoV-2 infection by a healthcare professional.
- Able to provide consent to participation in the study or with legal representative/parents that are able to consent to participation.
- Pregnant women.

<u>Exclusion Criteria</u>

- Not likely able to tolerate sample collection procedures.
- Unable to provide saliva and NPS Swab.
- Individuals with involvement in the planning and/or conduct of the study (company staff and/or site staff) individuals employed by the site or sponsor unless a registered patient at the study site.
- Individuals who have previously participated in a clinical study using the Sal6830™ SARS-CoV-2 Saliva Test

### 3) **Combined questionnaire result tables**

| | | | | | | |
|---|---|---|---|---|---|---|
| **CLINICAL TRIAL HEALTHCARE PROFESSIONAL: Letter and number codes consistent with manuscript text.** | SITE | The MicroGEM Sal 6830 SARS-CoV-2 Saliva Test works well in Point of Care settings | I found the MicroGEM Sal 6830 SARS-CoV-2 Saliva Test was unnecessarily complex | I found the MicroGEM Sal 6830 SARS-CoV-2 Saliva Tests easy to use | I will need the support of a technical person to be able to use the MicroGEM Sal 6830 SARS-CoV-2 Saliva Test | I found the various functions in the MicroGEM Sal 6830 SARS-CoV-2 Saliva Testwell integrated |
| **4** | Care 4 U | 4 | 1 | 4 | 1 | 4 |
| **1** | Care 4 U | 5 | 1 | 5 | 1 | 5 |
| **2** | Care 4 U | 5 | 1 | 5 | 1 | 5 |
| **3** | Care 4 U | 4 | 1 | 3 | 1 | 4 |
| **6** | Care 4 U | 5 | 2 | 5 | 2 | 5 |
| **7** | Care 4 U | 5 | 1 | 5 | 1 | 5 |
| **5** | Care 4 U | 5 | 1 | 5 | 1 | 5 |
| **B** | St. Mary’s | 4 | 2 | 4 | 1 | 4 |
| **A** | St. Mary’s | 5 | 1 | 5 | 1 | 5 |

| | | | | | | |
|---|---|---|---|---|---|---|
| **CLINICAL TRIAL HEALTHCARE PROFESSIONAL: Letter and number codes consistent with manuscript text.** | I thought there was inconsistency in the MicroGEM Sal 6830 SARS-CoV-2 Saliva Test | I would imagine that most healthcare professionals would learn to use the MicroGEM Sal 6830 SARS-CoV-2 Saliva Test quickly | I found the MicroGEM Sal 6830 SARS-CoV-2 Saliva Test difficult to use | I felt confident using the MicroGEM Sal 6830 SARS-CoV-2 Saliva Test | I needed to learn a lot of things before I could get going with the MicroGEM Sal 6830 SARS-CoV-2 Saliva Test | I found the MicroGEM Sal 6830 SARS-CoV-2 Saliva Test results easy to interpret |
| **4** | Not Asked | 4 | 1 | 5 | 3 | 5 |
| **1** | Not Asked | 4 | 2 | 5 | 5 | 5 |
| **2** | Not Asked | 5 | 1 | 5 | 1 | 5 |
| **3** | Not Asked | 4 | 1 | 5 | 1 | 5 |
| **6** | 1 | 5 | 1 | 4 | 2 | 5 |
| **7** | 1 | 5 | 1 | 5 | 1 | 5 |
| **5** | 1 | 5 | 1 | 5 | 1 | 5 |
| **B** | Not Asked | 5 | 1 | 5 | 1 | 5 |
| **A** | Not Asked | 5 | 1 | 5 | 1 | 5 |

| | |
|---|---|
| **CLINICAL TRIAL HEALTHCARE PROFESSIONAL: Letter and number codes consistent with manuscript text.** | Would you recommend the device to another (non laboratory) healthcare professional? |
| **4** | Definitely yes |
| **1** | Definitely yes |
| **2** | Definitely yes |
| **3** | Probably yes |
| **6** | Definitely yes |
| **7** | Definitely yes |
| **5** | Definitely yes |
| **B** | Definitely yes |
| **A** | Definitely yes |
